# Incidence of COVID-19 in a cohort of adult and paediatric patients with rheumatic diseases treated with targeted biologic and synthetic disease-modifying anti-rheumatic drugs

**DOI:** 10.1101/2020.04.30.20086090

**Authors:** Xabier Michelena, Helena Borrell, Mireia López-Corbeto, María López-Lasanta, Estefanía Moreno, María Pascual-Pastor, Alba Erra, Mayte Serrat, Esther Espartal, Susana Antón, Gustavo Adolfo Añez, Raquel Caparrós-Ruiz, Andrea Pluma, Ernesto Trallero-Araguás, Mireia Barceló-Bru, Miriam Almirall, Juan José De Agustín, Jordi Lladós Segura, Antonio Julià, Sara Marsal

**Affiliations:** Rheumatology Department, Rheumatology Research Group, Vall d’Hebron Research Institute, Vall d’Hebron University Hospital, Barcelona, Spain; Leeds Institute of Rheumatic and Musculoskeletal Medicine, University of Leeds, Leeds, United Kingdom

## Abstract

**OBJECTIVES:** To investigate the incidence of COVID-19 in a cohort of adult and paediatric patients with rheumatic diseases receiving targeted biologic and synthetic disease modifying anti-rheumatic drugs (tDMARDs) and to explore the possible effect of these treatments in the clinical expression of COVID-19.

**METHODS:** A cross-sectional study comprising of a telephone survey and electronic health records review was performed including all adult and paediatric patients with rheumatic diseases treated with tDMARDs in a large rheumatology tertiary centre in Barcelona, Spain. Demographics, disease activity, COVID-19 related symptoms and contact history data were obtained from the start of the 2020 pandemic. Cumulative incidence of confirmed cases (SARS-CoV-2 positive PCR test) was compared to the population estimates for the same city districts from a governmental COVID-19 health database. Suspected cases were defined following WHO criteria and compared to those without compatible symptoms.

**RESULTS:** 959 patients with rheumatic diseases treated with tDMARDs were included. We identified 11 confirmed SARS-CoV-2 positive cases in the adult cohort and no confirmed positive cases in the paediatric cohort. All patients had a successful recovery and only one patient required admission in the intensive care unit. When using the same classification criteria (only COVID-19 positive cases with pneumonia), COVID-19 incidence rates of the rheumatic patient cohort were very similar to that of the general population [(0.48% (95% CI 0.09 to 8.65%)] and [0.58% (95% CI 5.62 to 5.99%)], respectively. We found significant differences in tDMARDs proportions between the suspected and non-suspected cases (p=0.002).

**CONCLUSION:** Adult and paediatric patients with rheumatic diseases on tDMARDs do not seem to present a higher risk of COVID-19 or a more severe disease outcome when compared to general population. Our exploratory analysis suggests that the proportion of COVID-19 suspected cases differs between tDMARDs.

## INTRODUCTION

The Coronavirus 19 disease (COVID-19) caused by the SARS-CoV-2 virus has been declared a global health emergency in 2020. Health systems around the globe are struggling to find efficacious treatments against COVID-19 and the associated acute respiratory distress syndrome (ARDS). Previous experience with SARS and MERS-CoV infection demonstrated a marked pro-inflammatory response (Th1 and Th17) in patients with ARDS which is also seen in COVID-19 (1–3). This evidence has prompted the off-label use of IL-6 inhibitors in severe COVID-19 disease as well as the initiation of randomised clinical trials to test these drugs (clinicaltrials.gov, NCT04320615). Additionally, other inflammatory cytokine pathways such as the IL-1 pathway are currently being explored (clinicaltrials.gov, NCT04324021). Hydroxychloroquine, a commonly used antirheumatic medication, has been shown to have *in vitro* protective effects against SARS-CoV-2 infection (4), although its clinical efficacy still needs to be adequately assessed via randomized clinical trials (5). A recently approved RA drug, JAK1/2 inhibitor baricitinib, could also be protective by reducing the ability of the virus to infect lung cells (6). Therefore, patients with rheumatic diseases provide an opportunity to rapidly learn the impact of immunosuppressive agents as protective drugs against SARS-CoV-2 infection and against the development of more severe outcomes in COVID-19 disease.

Targeted biological and synthetic disease modifying antirheumatic drugs (tDMARDs) could also have a detrimental effect in COVID-19 disease. There is a major need in the rheumatological community to determine which, if any, tDMARDs increase the vulnerability to infection and should therefore be stopped. The European League Against Rheumatism (EULAR), American College of Rheumatology (ACR), National Institute for Health and Care Excellence (NICE) and the Paediatric Rheumatology European Society (PRES) have published preliminary guidance in this scenario (7–10). EULAR and PRES suggest that patients should discuss treatment discontinuation with their rheumatologists. ACR recommendations also favour to temporarily stop treatment with tDMARDs if exposure to SARS-COV-2. NICE and ACR advocate the suspension of tDMARDs if COVID-19 is confirmed or suspected with the ACR considering IL-6 inhibitors an exception, which may be continued under specific circumstances. So far, these recommendations are based on expert opinion as there is yet very limited epidemiological evidence on the potential risk conferred by tDMARDs with regards to severe COVID-19 disease complications in patients with rheumatic diseases. A recent report from a clinical centre in Pavia, one of the most affected regions in Italy, analysed 320 adult rheumatic patients and found no evidence of risk increase or decrease (11). To date, only one report from a paediatric centre in Milan shows no confirmed COVID-19 cases in children (12). With this scant evidence, rheumatologists worldwide are in the dark as to how to manage patients on immunosuppressive therapies during the pandemic and more information is urgently needed (13). The aim of our study was to report the incidence of COVID-19 in a cohort of adult and a cohort of paediatric patients with rheumatic diseases receiving tDMARDs from a reference tertiary hospital in Spain, and to explore the possible effect of these treatments in the expression of COVID-19.

## METHODS

### Study design

A cross-sectional study consisting of a telephone interview followed by a comprehensive review of electronic health records was conducted in a tertiary centre hospital in Barcelona, Spain, during the 2020 COVID-19 pandemic. All patients had a diagnosed inflammatory rheumatic disease treated with tDMARDs attending the Rheumatology Department in Vall d’Hebron University Hospital, Barcelona, Spain. Vall d’Hebron University Hospital is the largest hospital complex in the Catalonia region and one of the most important in Spain being a reference centre for adult and paediatric rheumatology.

### Study population

All adult and paediatric patients with the following clinical diagnosis: rheumatoid arthritis (RA), psoriatic arthritis (PsA), axial spondyloarthritis (axSpA), juvenile idiopathic arthritis (JIA) and autoinflammatory syndromes (AIS) who were receiving any of the following treatments: anti-TNF alpha drugs (etanercept, adalimumab, infliximab, golimumab and certolizumab), IL-1 inhibitors (anakinra), IL-6 inhibitors (tocilizumab and sarilumab), IL 12/23 inhibitors (ustekinumab), IL-17 inhibitors (secukinumab and ixekizumab), CTLA4-Ig (abatacept), JAK inhibitors (tofacitinib, baricitinib and ruxolitinib) and PDE4 inhibitors (apremilast) at the time of the study were invited to take part.

### Demographic, clinical and questionnaire data

The following variables were retrieved from the hospital electronic health record: age, sex, smoking status, comorbidities (obesity, hypertension, diabetes, previous lung disease, cardiovascular diseases, neoplasia), rheumatic disease diagnosis, current targeted immunosuppressive agent, concomitant treatment with methotrexate and hydroxychloroquine and present glucocorticoids dose. The most recent disease indexes (<4 months) encompassing the Disease Activity Score-28 (DAS-28) for RA, the Bath Ankylosing Spondylitis Disease Activity Index (BASDAI) for axSpA and the Disease Activity in PSoriatic Arthritis score (DAPSA) for PsA were included in the analysis.

The phone survey was performed by experienced professionals from the rheumatology department after adequate training during a 2-week period (March 26^th^-April 10^th^ 2020) after lockdown was implemented by the Spanish government on March 14^th^ 2020. COVID-19-related symptomatology as well as epidemiological and contact data since the start of the pandemic in the Barcelona area was obtained from all patients. The detailed questionnaire can be found in the supplementary material.

For this analysis, we considered “confirmed” cases when the SARS-CoV-2 polymerase chain reaction (PCR) was performed and resulted positive. Due to regional health policies(14), SARS-CoV-2 PCR testing was restricted to patients that had to be admitted to the hospital due to lung involvement. Additionally, clinical and radiological features, blood analysis results and outcome were investigated in patients with confirmed COVID-19 disease.

Following the World Health Organisation (WHO) guidance(15), we considered “suspected” cases when patients reported fever plus one other respiratory symptom (dyspnoea, persistent cough or odynophagia) OR presented 1 of the previous symptoms (fever, dyspnoea, persistent cough or odynophagia) and had had a contact with a confirmed or probable case.

### Statistical analysis

Data are presented for both adult and paediatric subjects, defining adults as subjects above 18 years old. Descriptive statistics were used to compare patient and disease characteristics according to classification as suspected cases. Association between disease activity and suspected cases was explored considering active disease when DAS28>2.6 in RA, BASDAI ≥4 in axSpA and DAPSA >14 in PsA. Pearson’s chi square, Fisher’s exact test, Student’s t-test and Wilcoxon rank sum test were used for comparisons as appropriate. A test for linear trend was used to explore the relationship between the level of contact exposure with the proportion of suspected cases.

Cumulative incidence was adjusted for sex and age by direct standardisation, using a COVID-19 epidemiological database from Barcelona generated by the local health institution(16). This database is updated daily and contains district-level information of COVID-19 confirmed cases. For this analysis we selected the data from the most frequent postal codes within our cohort, globally representing 50% of the patients. Cumulative confirmed cases were updated until April 10^th^ 2020. An additional analysis only considering confirmed cases who presented pneumonia in the chest x-ray hence requiring hospital admission (same testing criteria as for general population) was performed. Analyses were performed with STATA v 16.0 and RStudio v.3.5.1.

## RESULTS

A total of 1,045 patients with rheumatic diseases currently taking tDMARDs were contacted, and n=959 completed the survey. Following the WHO criteria, 95 patients were classified as suspected cases for COVID-19 infection, 11 of whom were subsequently confirmed to have a positive SARS-CoV-2 PCR test.

### Confirmed COVID-19 cases

Clinical characteristics of the 11 COVID-19 confirmed cases are summarised in Table 1. No confirmed cases were identified in the paediatric cohort. Patients had a median age of 45 years (IQR 30-63) and there was a 54.6% male prevalence. Briefly, the most commonly reported symptom was fever (82%), followed by persistent cough (73%) and malaise (55%). Six patients had radiographic findings of pneumonia on admission. All patients successfully recovered from COVID-19 disease after a median stay of 9.5 days (IQR 5-20 days) and only 1 patient required intensive unit care being the one with more comorbidities. The tDMARD was not administered during the admission in the cases with COVID-19 pneumonia, although in 50% of cases the usual treatment dose did not coincide with that timepoint. Of the remaining 5 cases who did not require admission, the tDMARD was maintained in 4 of them with further successful recovery and no disease flare. Most patients had a contact with a confirmed or suspected case either at home (2 patients had contact with a confirmed case and 5 patients with a suspected case), at work (2 patients had contact with a confirmed case and 4 patients with a suspected case) or both (1 patient).

**Table 1.**
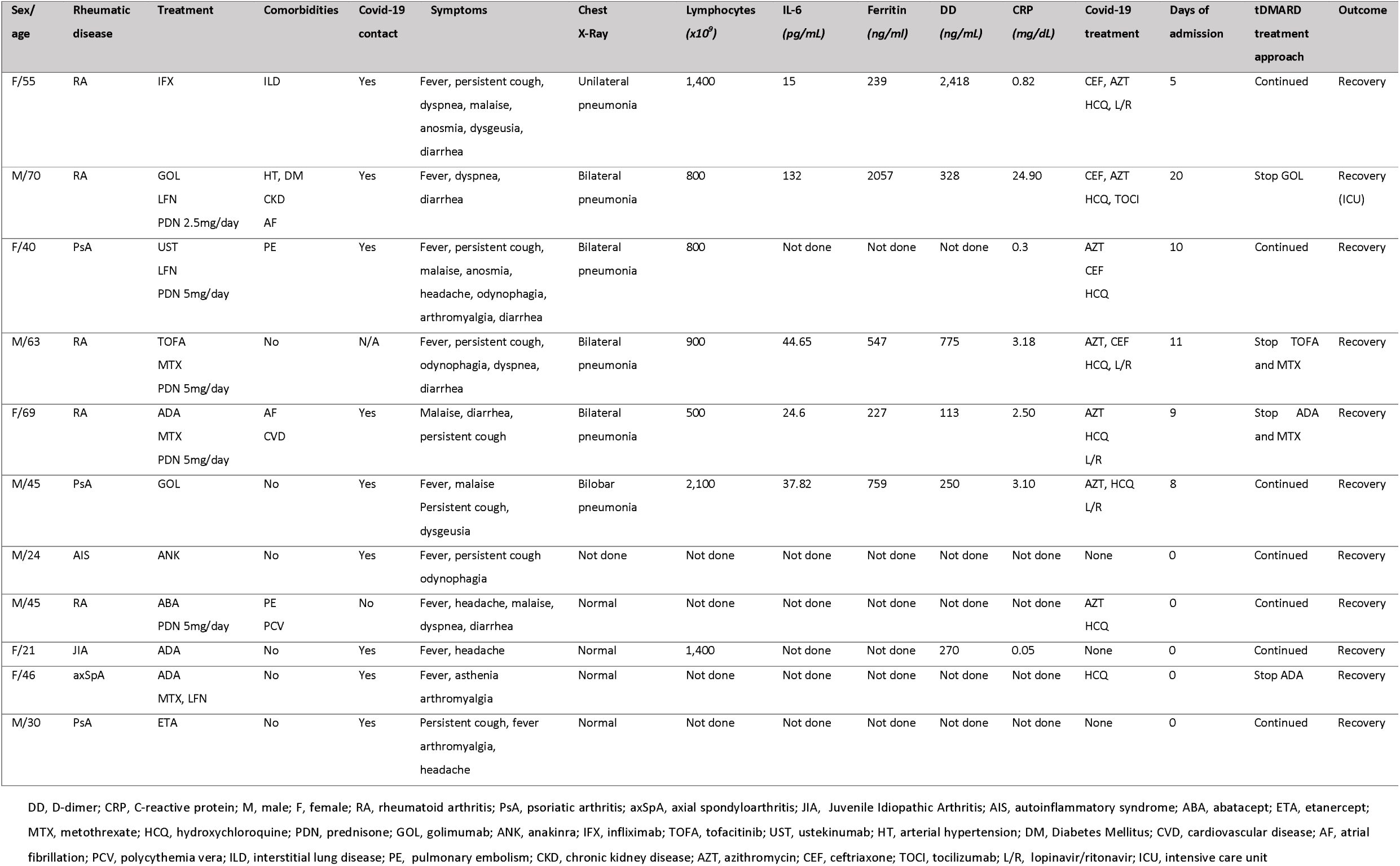
Confirmed cases of COVID-19 in tDMARD treated inflammatory rheumatic disease patients

### Cumulative incidence rates

Crude and adjusted incidence rates stratified by sex and age group are presented in Table 2. When adjusted for same age, sex and city districts, the cumulative incidence rate in our population was found to be 1.21% (95% CI 0.42-19.94%) compared to 0.58% (95% CI 5.62-5.99%) in the general population. If restricting to those confirmed cases with pneumonia, the adjusted cumulative incidence rate of rheumatic patients was found to be more similar to that of the general population [0.48% (95% CI 0.09-8.65%) vs 0.58% (95% CI 5.62-5.99%)].

**Table 2.**
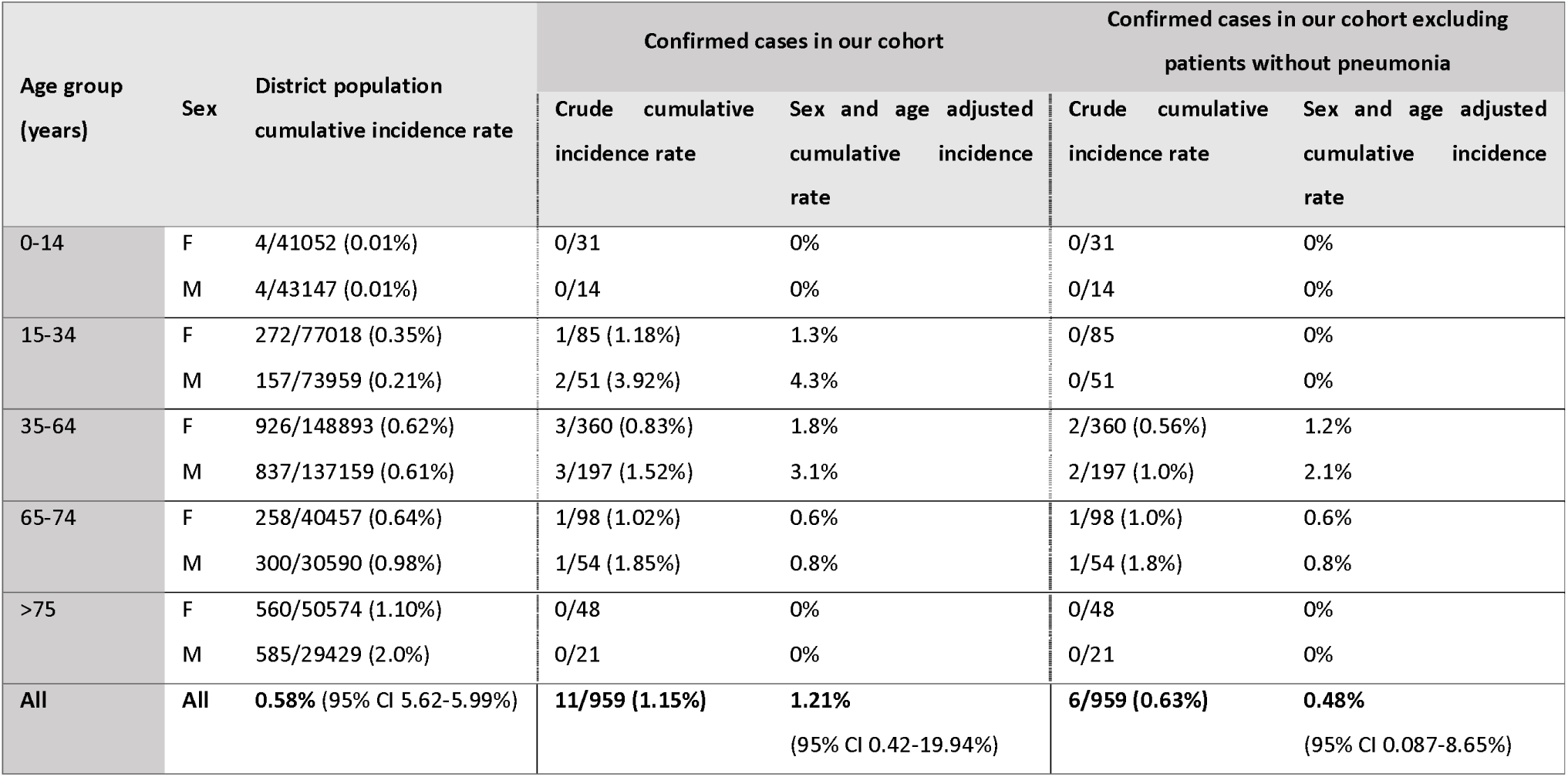
Cumulative incidence rates of COVID-19 confirmed cases in our cohort and selected sample population (crude and adjusted)

### Suspected vs. non-suspected cases

Comparing the clinical characteristics of the suspected cases in the adult and paediatric cohorts with those of unaffected patients without these symptoms (Table 3), adult patients with compatible symptoms were younger (45.7 vs. 54.2 years, p<0.001). A total of 8 suspected cases were documented in the paediatric group. When exploring the effect of therapies between groups (suspected vs. non-suspected), we found a significant association in adults (p=0.002). CTLA-4-Ig and anti-IL6 therapies showed the lowest incidence of COVID-19-compatible symptoms (<5% at the treatment level) among adult rheumatic patients. In the paediatric population, all suspected e cases were treated with anti-TNF therapy. However, when comparing to the adult anti-TNF treated sub-group we found no significant difference in risk (p=0.33).

**Table 3.**
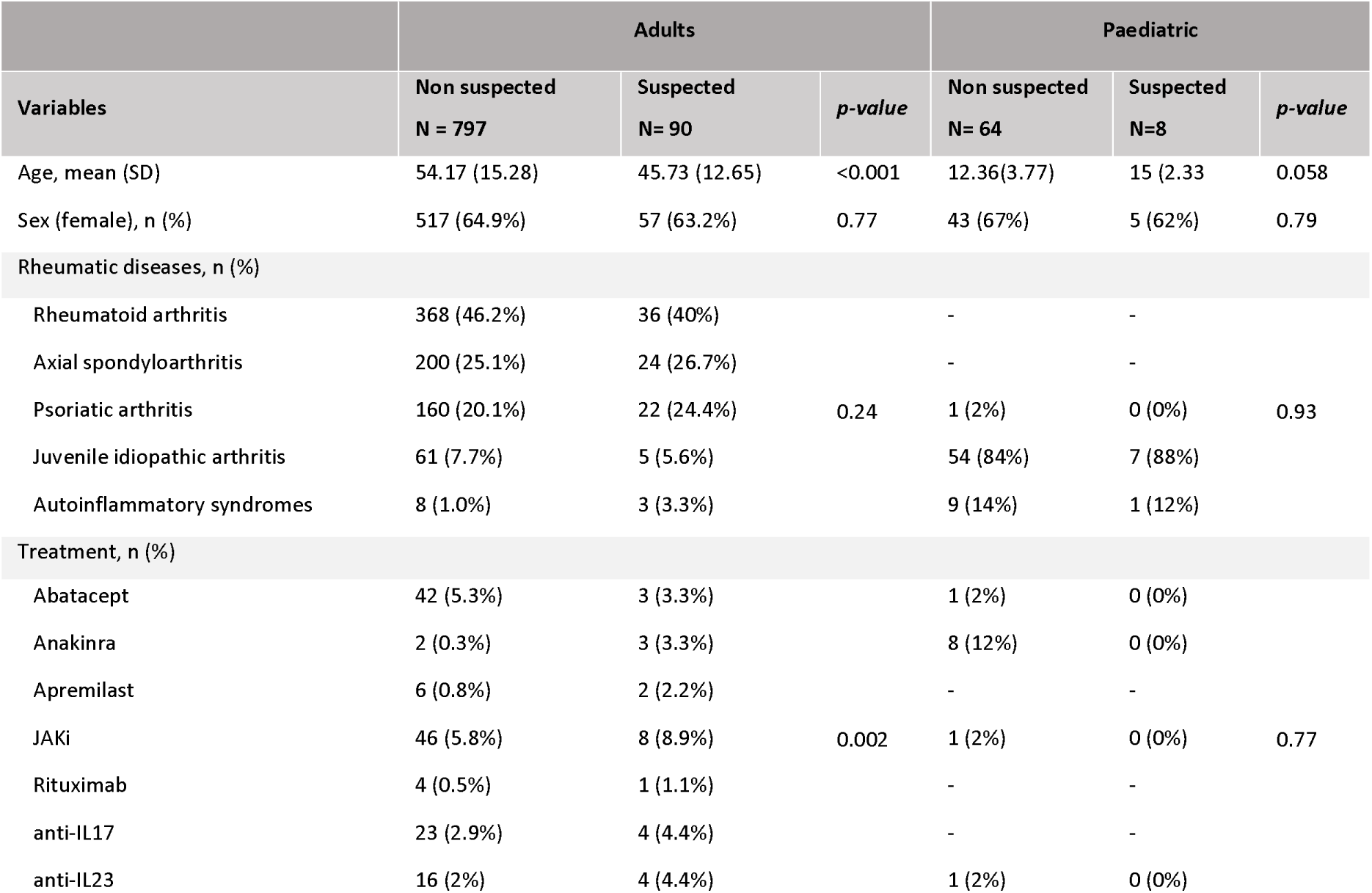
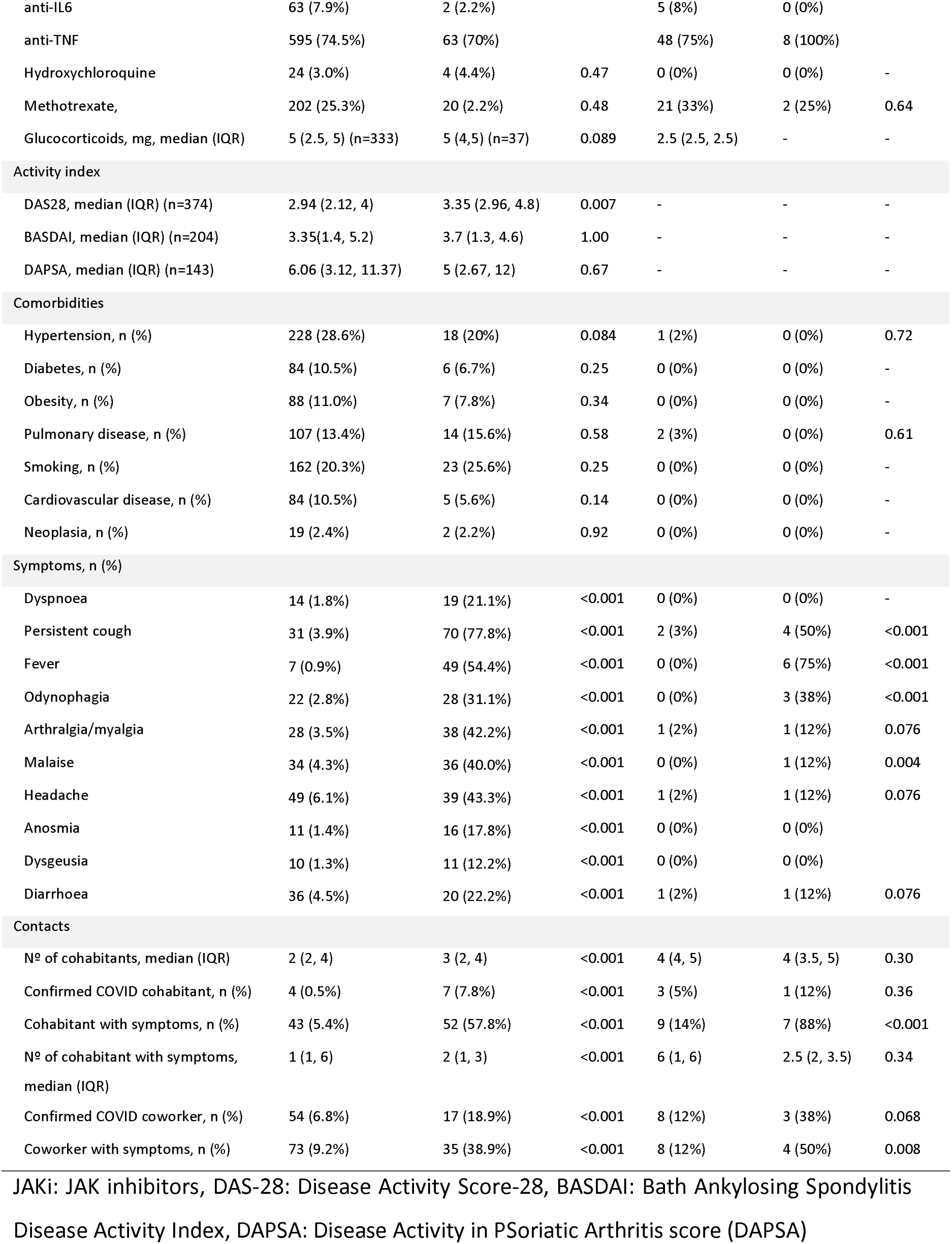
Suspected and non-suspected case characteristics in adult and paediatric population

As expected, in both adult and paediatric patients, all symptoms were significantly more frequent in the suspected case group and patients had a higher exposure to possible sources of infection with more number of cohabitants (trend test p<0.001) with confirmed COVID-19, cohabitants with compatible symptoms, number of cohabitants with compatible symptoms (trend test p <0.001) and co-workers with confirmed disease or compatible symptoms.

### Disease activity

RA patients with active disease (DAS28>2.6) were more likely to be classified as suspected case [10.8% active vs 3.5% non-active, diff: −7.8% (CI 95% −12.2, −2.2%) p= 0.012], Active RA patients were older (59.9 vs 56.3 years, p=0.010) and had more prevalence of diabetes (14.2% vs 7.0%, p=0.035), obesity (12.1% vs. 5.6, p=0.041) and current prednisone dose (median dose: 5 vs 4.5 mg, p=0.003). There were no differences in proportion of active disease patients comparing suspected and non-suspected cases in the axSpA [11.8% BASDAI active vs 10.2% BASDAI non-active, diff −1.6% (CI-10.6, 7.2), p=0.71] or PsA groups [8.0% DAPSA active vs 12.7% DAPSA non-active, diff: 4.7% (CI 95% −7.5, 16.9%) p= 0.51].

## DISCUSSION

Spain is one of the countries in the world where the SARS-CoV-2 expanded more rapidly on the first trimester of 2020. People receiving immunosuppressant therapies are considered more susceptible to viral and bacterial infections and, consequently, a major concern for rheumatologists worldwide is to know if rheumatic patients receiving these therapies have an increased risk of COVID-19. In this study, we performed an epidemiological survey of the COVID-19 incidence in rheumatic patients receiving tDMARD therapy from a large tertiary hospital from Barcelona, Spain. From a total of 959 IMID patients treated with tDMARDs, we identified 11 confirmed COVID-19 cases in adults and no cases in the paediatric population. When comparing our cohort to the epidemiological data from the same city districts and tested with the same criteria, we found that the cumulative incidence for rheumatic patients is highly similar to that of the general population. In our patient cohort, 10% of patients had COVID-19 compatible symptoms according to the WHO criteria. Analysing the tDMARD distribution in these at-risk cases, we found significant differences in tDMARDs proportions between the suspected and non-suspected cases. To our knowledge, this is the largest study outlining the extent of COVID-19 in rheumatic patients treated with tDMARDs to date.

Importantly, this study provides additional evidence that rheumatic patients on immunosuppressant therapies are not at a higher risk of COVID-19 infection than the general population. The number of confirmed cases in our cohort is comparable to that recently reported from one of the focal regions in Italy (1.2% vs. 1.2%)(11). If considering all cases, the adjusted incidence rate appears to be higher in our patients compared to the general population (1.21% vs 0.58%). However, these results need to be contextualised. Following hospital and regional health guidance(14), SARS-CoV-2 PCR confirmation should be only performed in patients with pneumonia and hospital admission requirements. In our cohort, however, 5 additional patients with normal chest x-ray and without inpatient admission criteria were PCR-tested for SARS-CoV-2 PCR. These patients were health workers from the same hospital (n=2) or patients who were tested as considered to be vulnerable population by the treating clinician (n=3). Once these subjects are excluded from the analysis, the incidence rate is analogous to the population from the same city area. Reassuringly, tDMARD-treated inflammatory rheumatic disease patients do not seem to present a more severe clinical presentation of COVID-19, although our sample of confirmed cases is small. All of the confirmed cases had a complete recovery and only one required admittance in the intensive care unit. Of relevance, there were no confirmed cases in our paediatric cohort as shown in a previous report from Italy (12). This result is in concordance with the low occurrence of COVID-19 reported for children from the general population (17).

At present, as outlined in the introduction EULAR, ACR, NICE and PRES guidance regarding patient management during the COVID-19 pandemic propose individualised decisions on tDMARDs (7–10). ACR and NICE also advocate for a tDMARDs stop if COVID-19 is suspected or confirmed. In our rheumatology department, tDMARD treatment discontinuation was performed individually, based on the specific patient characteristics. Of note, in most of the confirmed cases that were not admitted (i.e. normal chest x-ray), treatment was not stopped and all patients completely recovered from the infection. This is in line with the recently published guidelines of the German Society of Rheumatology that in a situation of a positive SARS-CoV-2 PCR without signs of infection, only recommend a tDMARD pause or delay for 5-6 days(18). Altogether, our data supports a tDMARD maintenance strategy during the pandemic, although more evidence from different registries is needed.

Similar to recent reports (19), we found that most COVID-19 infected cases do not require secondary care. This situation clearly hinders the estimation of the true population incidence. In our study, to better explore the clinical and epidemiologic characteristics of the suspected cases we decided to identify which patients had compatible symptoms based on the current WHO guidance (15). The most frequent symptoms in both confirmed and suspected cases were fever and persistent cough as outlined in a recent meta-analysis (20). Patients with compatible symptoms were significantly younger in the adult population. Factors that could contribute to this increase could be that younger patients tend to live with a higher number of cohabitants and are more likely to leave home for work and, therefore, have an increased risk of infection exposure. We found no differences in comorbidities between groups. This is in apparent contradiction to data suggesting that comorbidities have an important role in COVID-19 severity (21). However, it is possible that comorbidities influence the severity of the disease rather than the probability of infection (22). Finally, in our cohort active RA patients were more likely to be classified as suspected cases. There is evidence that RA patients are at an increased risk of infections particularly if they present high disease activity and depending on the concomitant glucocorticoids dose (23–25). Also, when exploring the characteristics of these active patients, we found they were older, more obese and had more diabetes suggesting an effect of comorbidity in this finding (26).

Although treatment data should be interpreted with caution, we identified significant differences in tDMARDs proportions between the suspected and non-suspected cases. Suspected cases were less likely to be treated with abatacept or IL-6 inhibitors than with the other therapies (3.6% vs 10.7%, respectively, p=0.016). This data suggests a protective effect of these two therapies. IL-6 is one of the cytokines more highly expressed in severe forms of COVID-19, and has been associated to the reported cytokine storms (27). For this reason, IL-6 inhibitors are currently being used in the treatment of severe COVID disease (28). Our results support the protective role of IL-6 inhibition in the progression of disease severity and suggest this could be applied preventively, at earlier stages of the infection (29). There is preliminary evidence that inhibition of the CD80/86 co-stimulation by abatacept as well as IL-6 inhibition could be useful to treat interstitial lung disease (30, 31). This potential protective effect on the target organ of inflammation in COVID-19 could explain the reduced risk observed in rheumatic patients. Moreover, abatacept has been associated with a lower risk of serious infections which adds up as a possible explanation of these findings (32). Nonetheless, randomised controlled trials are needed to corroborate the protective effect of these treatments in COVID-19.

Some limitations of this study need to be addressed. First, we designed an observational study based on a telephone survey with different date completion and filled by distinct operators. Patient reporting could have been influenced by the operator and symptoms could have been missed as these could have appeared after the date of completion. To address this potential bias, we reviewed the electronic health records of all patients to detect possible cases and asked all our patients to actively contact us if any of the compatible symptoms occurred. Secondly, we only included rheumatic patients treated with tDMARDs without a control group, either treated with other type of DMARDs or untreated patients. Thirdly, our study is mainly focused on chronic inflammatory arthritis and we did not include, for example, patients with systemic lupus erythematosus or giant cell arteritis that could provide additional insights into the protective effect of hydroxychloroquine and anti-IL6 therapies. Further, the lockdown effect might be more pronounced in IMID patients since they are considered an immunosuppressed cohort protecting them from possible exposure to infection and, therefore, distorting their comparison with general population. With regards to the analysis, the small number of cases did not allow for the adjustment for potential confounders other than age and sex. Lastly, due to limited availability of SARS-CoV-2 testing in our setting, the estimate of COVID-19 incidence is inaccurate, thereby prompting the health community to rely on suspect cases definitions that might not precisely identify all cases.

In conclusion, adult and paediatric patients with rheumatic diseases treated with tDMARDs do not seem to present a higher risk of COVID-19 or a more severe disease outcome compared to the general population. These data suggest that tDMARDs should not be stopped in these patients during the pandemic. Our study also suggests that tDMARDs show different levels of protection against COVID-19 and supports the development of randomized clinical trials to adequately assess their individual effect. International initiatives currently under way, such as the Global Rheumatology Alliance and the EULAR COVID-19 database will provide invaluable data for rheumatic disease management, but epidemiological studies from large national reference centres can be extremely helpful as an interim guidance (13, 33). In this unprecedented time, when therapies commonly used in rheumatic diseases could prove useful to manage a pandemic disease, rheumatologists are required to provide all their expertise to accelerate the generation of scientific data to protect the lives of people, not only those affected by rheumatic diseases, but also for the global population.

## Data Availability

Data is available upon request to corresponding author.

## Acknowledgements

We thank Dr Helena Marzo-Ortega for critical review of the final manuscript. The authors would also like to thank Elena Granell and Montserrat Sender for their help in the survey coordination. A special thank you to all patients who contributed with their time to participate in the study in these difficult times.

## Contributors

XM, HB and SM contributed to the conception and design of the study. XM, HB, MLC, MLL, EM, MPP, AED, MSL, EE, SA, GAS, RCR, APS, ETA, MBB, MAB, JDD and JL were involved in data collection. XM and AJ performed the data analysis and interpretation of data. All authors contributed to drafting and/or revising the manuscript.

## Competing interests

XM received speaker fees from Novartis and Sanofi-Genzyme. HB received speaker fees from Lilly, MSD and Pfizer. RCR received speaker fees from MSD. ETA received speaker fees from Roche and Bristol-Myer-Squib outside this work. SM received speaker fees, grant and honoraria from Roche and Bristol-Myer-Squib outside this work. MLC, MLL, EM, MPP, AED, MSL, EE, SA, GAS, APS, MBB, MAB, JDD, JL and AJ have nothing to disclose.

### Patient consent

Obtained.

### Ethics approval

The study was approved by the Hospital Universitari Vall d’Hebron Clinical Research Ethics Committee (No 5633). This study was conducted according to the principles of the Declaration of Helsinki.

## Patient and Public Involvement

Patients or the public were not involved in the design, or conduct, or reporting, or dissemination plans of our research.

## References

1. Wong CK, Lam CW, Wu AK, Ip WK, Lee NL, Chan IH, et al. Plasma inflammatory cytokines and chemokines in severe acute respiratory syndrome. Clin Exp Immunol. 2004;136(1):95–103.

2. Mahallawi WH, Khabour OF, Zhang Q, Makhdoum HM, Suliman BA. MERS-CoV infection in humans is associated with a pro-inflammatory Th1 and Th17 cytokine profile. Cytokine. 2018;104:8–13.

3. Huang C, Wang Y, Li X, Ren L, Zhao J, Hu Y, et al. Clinical features of patients infected with 2019 novel coronavirus in Wuhan, China. Lancet. 2020;395(10223):497–506.

4. Liu J, Cao R, Xu M, Wang X, Zhang H, Hu H, et al. Hydroxychloroquine, a less toxic derivative of chloroquine, is effective in inhibiting SARS-CoV-2 infection in vitro. Cell Discov. 2020;6:16.

5. Ferner RE, Aronson JK. Chloroquine and hydroxychloroquine in covid-19. BMJ. 2020;369:m1432.

6. Richardson P, Griffin I, Tucker C, Smith D, Oechsle O, Phelan A, et al. Baricitinib as potential treatment for 2019-nCoV acute respiratory disease. Lancet. 2020;395(10223):e30-e1.

7. European League Against Rheumatism (EULAR). EULAR Guidance for patients COVID-19 outbreak 2020 [updated 17/03/2020; cited 28/04/2020]. Available from: https://www.eular.org/eularguidanceforpatientscovid19outbreak.cfm.

8. American College of Rheumatology (ACR). COVID-19 Clinical Guidance for Adult Patients with Rheumatic Diseases 2020 [updated 14/04/2020; cited 28/04/2020]. Available from: https://www.rheumatology.org/Announcements#ClinicalGuidance.

9. National Institute for Health and Care Excellence (NICE). COVID-19 rapid guideline: rheumatological autoimmune, inflammatory and metabolic bone disorders (NG167) 2020 [updated 24/04/2020; cited 28/04/2020]. Available from: https://www.nice.org.uk/guidance/ng167.

10. Paediatric Rheumatology European Association (PRES). PRES recommendations for coronavirus outbreak 2020 [updated 16/03/2020; cited 28/04/2020]. Available from: https://www.pres.eu/news/newsstory.html?id=29.

11. Monti S, Balduzzi S, Delvino P, Bellis E, Quadrelli VS, Montecucco C. Clinical course of COVID-19 in a series of patients with chronic arthritis treated with immunosuppressive targeted therapies. Ann Rheum Dis. 2020 published on 2020/04/04. doi:10.1136/annrheumdis-2020-217424.

12. Filocamo G, Minoia F, Carbogno S, Costi S, Romano M, Cimaz R. Absence of severe complications from SARS-CoV-2 infection in children with rheumatic diseases treated with biologic drugs. J Rheumatol. 2020 published on 27/04/2020. doi:10.3899/jrheum.200483.

13. Robinson PC, Yazdany J. The COVID-19 Global Rheumatology Alliance: collecting data in a pandemic. Nat Rev Rheumatol. 2020 published on 2020/04/04. doi:10.1038/s41584-020-0418-0.

14. Sub-direcció General de Vigilància i Resposta a Emergències de Salut Pública. Procediment d’actuació enfront de casos d’infecció pel nou coronavirus SARS-CoV-2 2020 [updated 02/04/2020; cited 28/04/2020]. Available from: http://canalsalut.gencat.cat/coronavirus.

15. World Health Organisation (WHO). Global surveillance for COVID-19 caused by human infection with COVID-19 virus: interim guidance 2020 [updated 20/03/2020; cited 28/04/2020]. Available from: https://www.who.int/emergencies/diseases/novel-coronavirus-2019/technical-guidance/surveillance-and-case-definitions.

16. Agènda de Salut Pública de Barcelona (ASPB). Dades diàries de la infecció per coronavirus SARS-CoV-2 (COVID-19) a Barcelona 2020 [updated 10/04/2020; cited 28/04/2020]. Available from: https://www.aspb.cat/docs/COVID19aldiaBCN/.

17. Ludvigsson JF. Systematic review of COVID-19 in children shows milder cases and a better prognosis than adults. Acta Paediatr. 2020 published on 2020/03/24. doi:10.1111/apa.15270.

18. Schulze-Koops H, Specker C, Iking-Konert C, Holle J, Moosig F, Krueger K. Preliminary recommendations of the German Society of Rheumatology (DGRh eV) for the management of patients with inflammatory rheumatic diseases during the SARS-CoV-2/Covid-19 pandemic. Ann Rheum Dis. 2020 published on 2020/04/30. doi:10.1136/annrheumdis-2020-217628.

19. Wang Y, Wang Y, Chen Y, Qin Q. Unique epidemiological and clinical features of the emerging 2019 novel coronavirus pneumonia (COVID-19) implicate special control measures. J Med Virol. 2020 published on 2020/03/07. doi:10.1002/jmv.25748.

20. Fu L, Wang B, Yuan T, Chen X, Ao Y, Fitzpatrick T, et al. Clinical characteristics of coronavirus disease 2019 (COVID-19) in China: a systematic review and meta-analysis. J Infect. 2020. doi:10.1016/j.jinf.2020.03.041.

21. Yang J, Zheng Y, Gou X, Pu K, Chen Z, Guo Q, et al. Prevalence of comorbidities in the novel Wuhan coronavirus (COVID-19) infection: a systematic review and meta-analysis. Int J Infect Dis. 2020 published on 2020/03/17. doi: 10.1016/j.ijid.2020.03.017.

22. Preliminary Estimates of the Prevalence of Selected Underlying Health Conditions Among Patients with Coronavirus Disease 2019 - United States, February 12-March 28, 2020. MMWR Morb Mortal Wkly Rep. 2020;69(13):382–6.

23. Doran MF, Crowson CS, Pond GR, O’Fallon WM, Gabriel SE. Frequency of infection in patients with rheumatoid arthritis compared with controls: a population-based study. Arthritis Rheum. 2002;46(9):2287–93.

24. Au K, Reed G, Curtis JR, Kremer JM, Greenberg JD, Strand V, et al. High disease activity is associated with an increased risk of infection in patients with rheumatoid arthritis. Ann Rheum Dis. 2011;70(5):785–91.

25. Dixon WG, Abrahamowicz M, Beauchamp ME, Ray DW, Bernatsky S, Suissa S, et al. Immediate and delayed impact of oral glucocorticoid therapy on risk of serious infection in older patients with rheumatoid arthritis: a nested case-control analysis. Ann Rheum Dis. 2012;71(7):1128–33.

26. Lighter J, Phillips M, Hochman S, Sterling S, Johnson D, Francois F, et al. Obesity in patients younger than 60 years is a risk factor for Covid-19 hospital admission. Clin Infect Dis. 2020 published on 2020/04/10. doi:10.1093/cid/ciaa415.

27. Ye Q, Wang B, Mao J. Cytokine Storm in COVID-19 and Treatment. J Infect. 2020 published on 2020/04/14. doi:10.1016/j.jinf.2020.03.037.

28. Fu B, Xu X, Wei H. Why tocilizumab could be an effective treatment for severe COVID-19? J Transl Med. 2020;18(1):164.

29. McGonagle D, Sharif K, O’Regan A, Bridgewood C. The Role of Cytokines including Interleukin-6 in COVID-19 induced Pneumonia and Macrophage Activation Syndrome-Like Disease. Autoimmun Rev. 2020 published on 2020/04/07. doi: 10.1016/j.autrev.2020.102537:102537.

30. Manfredi A, Cassone G, Furini F, Gremese E, Venerito V, Atzeni F, et al. Tocilizumab therapy in rheumatoid arthritis with interstitial lung disease: a multicenter retrospective study. Intern Med J. 2019 published on 2019/10/30. doi:10.1111/imj.14670.

31. Fernandez-Diaz C, Loricera J, Castaneda S, Lopez-Mejias R, Ojeda-Garcia C, Olive A, et al. Abatacept in patients with rheumatoid arthritis and interstitial lung disease: A national multicenter study of 63 patients. Semin Arthritis Rheum. 2018;48(1):22–7.

32. Singh JA, Wells GA, Christensen R, Tanjong Ghogomu E, Maxwell L, Macdonald JK, et al. Adverse effects of biologics: a network meta-analysis and Cochrane overview. Cochrane Database Syst Rev. 2011;2011(2):Cd008794.

33. European League Against Rheumatism (EULAR). EULAR COVID-19 database 2020 [cited. Available from: https://www.eular.org/eularcovid19database.cfm.

